# Unmasking Hormonal Mechanisms of Hypertension in Obesity

**DOI:** 10.1101/2025.11.18.25340514

**Authors:** Stéfanie Parisien-La Salle, Cheng-Hsuan Tsai, Andrew J. Newman, Justin M. Chan, Julia Milks, Gail Adler, Arnaldo Ferrebus, Isabelle Hanna, Sanan Mahrokhian, Bertram Pitt, Jenifer M. Brown, Anand Vaidya

**Author notes:** Corresponding Author: Anand Vaidya, MD MMSc, Center for Adrenal Disorders, Mass General Brigham, Harvard Medical School, Boston, MA, 02115,.

## Abstract

**Background:** Understanding hormonal mechanisms of obesity-related hypertension may inform the optimal approach to targeted therapy.

**Objectives:** To interrogate hormonal phenotypes of obesity-related hypertension.

**Methods:** 77 participants with obesity and hypertension underwent deep-phenotyping of adrenocortical hormones at baseline and following multiple dynamic maneuvers to suppress and stimulate hormone production, including: the seated saline suppression (SST), oral sodium loading (OSLT) test, dexamethasone suppression test (DST), and ACTH-stimulation (ACTHstim). Participants were classified into 3 aldosteronism phenotypes: (1) primary aldosteronism (PA) phenotype (low renin with non-suppressible aldosterone), low-renin phenotype (low renin and low aldosterone), and renin-dependent aldosteronism (high renin with renin-mediated aldosteronism). The DST was used to evaluate for ACTH-independent hypercortisolism and ACTHstim was used to evaluate ACTH-modulated adrenocortical hormone production.

**Results:** Participants were 55.4 ± 9.4 years, 66.2% women, and had a BMI of 34.8 ± 5.2 kg/m^2^. At baseline, 37.7% of participants had a PA phenotype. Following sodium loading with SST, a persistent PA phenotype was seen in 28.5% and unmasked in an additional 23.4% of participants (total 51.9%). Participants with an unmasked PA phenotype had renin-dependent aldosteronism prior to SST, and thus were not identified during baseline testing. Persistent renin-dependent aldosteronism following SST was identified in 23.4% of the cohort and was characterized by greater kaliuresis and higher aldosterone levels (at baseline and following dynamic maneuvers to modulate ACTH and angiotensin II). These patterns were all reproduced following sodium loading with the OSLT. The DST identified ACTH-independent hypercortisolism in 9.2% of participants.

**Conclusions:** Over 80% of participants with obesity-related hypertension reproducibly exhibited pathologic phenotypes of aldosteronism and/or hypercortisolism. These overlapping hormonal mechanisms reveal the multi-factorial nature of obesity-related hypertension and provide evidence to support aldosterone- and cortisol-directed therapies to treat hypertension in people with obesity.

**CONDENSED ABSTRACT:** Understanding hormonal mechanisms of obesity-related hypertension may inform targeted therapy. Participants with obesity and hypertension underwent deep-phenotyping procedures to detect the: primary aldosteronism (PA) phenotype, low-renin phenotype, renin-dependent aldosteronism phenotype, and ACTH-independent hypercortisolism. In total, 51.9% of participants had a PA phenotype, of which approximately half also had superimposed renin-dependent aldosteronism. Another 23.4% participants had just renin-dependent aldosteronism that was characterized by higher aldosterone levels and kaliuresis, and 9.2% of participants had hypercortisolism. Over 80% of individuals with obesity-related hypertension exhibited overlapping pathologic phenotypes of aldosteronism and/or hypercortisolism, providing mechanistic evidence to support the efficacy of aldosterone- and cortisol-directed therapy (**Central Illustration**).

## INTRODUCTION

Over 70% of adults in the United States are either overweight or obese (1–3). Excess adiposity contributes to a range of diseases, including hypertension, diabetes, and dyslipidemia, which in turn drive cardiovascular disease and mortality, accounting for an estimated 5 million deaths and 160 million years of healthy life lost worldwide (4, 5).

Obesity is a major determinant in the development of hypertension, with epidemiologic data suggesting that excess adiposity accounts for 65-75% of the risk for essential hypertension (6). The link between adiposity and elevated blood pressure has been evident for decades (7, 8). Hypertension pathophysiology in obesity is complex and multifactorial, involving multiple neurohormonal mechanisms including dysregulation of the renin-angiotensin-aldosterone system (RAAS), cortisol dysregulation, sympathetic nervous system activation, renovascular disease, hypothalamic biology, and the adipose tissue secretome (9–13). Disentangling these overlapping mechanisms contributing to obesity-mediated hypertension could substantially inform the clinical care of hypertension and cardiovascular risk management as novel medical therapies addressing hormonal mediators of hypertension emerge (14–18). Herein, we conducted deep-phenotyping studies to interrogate hormonal phenotypes of obesity-related hypertension.

## METHODS

### Study Overview

Participants in this study were part of a prospective study that used a deep-phenotyping protocol to examine hormonal mechanisms involved in obesity-related hypertension (NCT04519164) (19). Herein, we conducted secondary analyses of the phenotyping studies to characterize hormonal mechanisms of obesity-related hypertension. This study was approved and monitored by the Mass General Brigham Human Research Institutional Research Board.

### Study Participants

Study participants were recruited from the local Boston, MA metropolitan area using a combination of social media advertisements and patient portal research invitations.

Participants were overweight or obese adults with hypertension (treated or untreated), elevated cardiovascular risk, and without diabetes. Participants were eligible for inclusion if they met one of two primary criteria: body mass index (BMI) ≥ 30 kg/m² with at least one of the following comorbidities, or BMI ≥ 25 kg/m² with at least two of the following comorbidities: (1) blood pressure ranging from 120–159/80–99 mmHg managed with zero to one antihypertensive agent; (2) dysglycemia (impaired fasting glucose 100–125 mg/dL or glycated hemoglobin 5.7–6.4%); (3) dyslipidemia (fasting triglycerides >150 mg/dL and HDL cholesterol <40 mg/dL in men or <50 mg/dL in women). Participants were excluded if they had a known diagnosis of primary aldosteronism (PA), abnormal kidney function (estimated glomerular filtration rate <60 mL/min/1.73 m²), serum potassium ≥5.3 mEq/L, a known diagnosis or treatment for type 1 or type 2 diabetes, a history of cardiovascular, ischemic changes on electrocardiogram, pregnancy or breastfeeding, or reported use of mineralocorticoid receptor antagonists (MRAs). In total, 80 participants were enrolled in the study.

### Study Protocol

Participants taking medications including diuretics, ACE inhibitors, and angiotensin receptor blockers were required to withdraw these drugs for two weeks prior to phenotyping procedures. During the washout period, if blood pressure exceeded 130/80 mmHg, doxazosin (2–8 mg/day) and/or amlodipine (2.5–10 mg/day) were initiated and titrated to an ideal target blood pressure of <130/80 mmHg using a combination of home and in-person blood pressure monitoring. After completing the medication withdrawal, participants completed 5 phenotypic protocols or maneuvers in the span of 2-3 weeks:

#### 1) Seated Saline Suppression Test

Participants underwent a standard seated, fasting saline suppression test (SST) beginning between 8:00 and 9:00 AM. This maneuver is an internationally recognized method for evaluating renin-independent aldosteronism, a hallmark of PA pathophysiology, and allows for comprehensive assessment of renin-angiotensin-aldosterone system dynamics (20). Saline infusion is used to induce intravascular volume expansion and physiologic suppression of renin and angiotensin II, thereby maximally suppressing angiotensin II-mediated aldosterone production. After maintaining an upright seated posture for 30 minutes, a baseline blood draw was performed, followed by 4 hours of seated posture while receiving two liters of intravenous infusion of normal saline (500 mL/h for 4 hours). Plasma aldosterone concentration (PAC) and plasma renin activity (PRA) were measured again at the end of the infusion. In total 77 of the enrolled participants completed the SST and had complete data for analysis.

#### 2) Oral Sodium Loading Test

Participants completed an oral sodium loading test (OSLT), a well-established maneuver to evaluate for PA pathophysiology that further permits interrogation of renin and aldosterone physiology (20). As with the SST, oral sodium loading is used to induce extracellular and intravascular volume expansion and a physiologic suppression of renin and angiotensin II, thereby maximally suppressing angiotensin II-mediated aldosterone production. Participants consumed 5 to 7 days of high dietary sodium diet (>200 mmol/d of sodium, 50 mmol/d of potassium and 600 mg/d of calcium), prepared in conjunction with professional dietary staff in the metabolic kitchen. After the last dietary day, participants presented fasting at 8:00 AM to the ambulatory clinical research center.

Participants performed a 24-hour urine collection prior to arrival. Seated blood draw was performed to measure PRA and PAC, and urine was used to measure 24h urine sodium and aldosterone. Of the 77 participants who completed the SST with available data, 65 also successfully achieved a 24h urine sodium of ≥180 mEq during the OSLT and were analyzed.

#### 3) Dexamethasone Suppression Test

To assess ACTH-independent cortisol and aldosterone production, participants underwent a dexamethasone suppression test (DST). Each participant was administered a 1 mg oral dose of dexamethasone between 11:00 PM and midnight. The following morning, participants arrived at the clinical research center for a fasting blood draw between 8:00 and 9:00 AM.

#### 4) ACTH Stimulation Test

Participants received a 250 mcg intravenous bolus of cosyntropin (synthetic ACTH peptide) to conduct an ACTH stimulation (ACTHstim) test, with a subsequent blood draw one hour later. This induces a physiologic stimulation of cortisol and aldosterone production.

#### 5) Blood Pressure Phenotyping

Screening blood pressure was measured after 5 minutes of seated rest, in triplicate, following standardized, automated, unattended, akin to the SPRINT trial (21).

Participants also completed 24h ambulatory blood pressure monitoring on *ad libitum* dietary intake after medication washout (Microlife WatchBP O3 Ambulatory; Microlife USA, Clearwater, FL) with automated measurements every 20 minutes from 07:00 AM to 10:00 PM and every 30 minutes from 10:00 PM to 06:00 AM. Seated blood pressure measurements were also taken during phenotyping visits.

### Phenotypic Classifications

Participants were classified into phenotypic categories based on renin and aldosterone dynamics at baseline and following sodium loading maneuvers. These classifications represented hormonal phenotypes rather than clinical diagnoses, and were evaluated before and after SST and OSLT to assess reproducibility and physiological relevance:

*Renin-Independent Phenotypes*:

- PA Phenotype: Characterized by overt renin-independent aldosterone production as defined as:

- At baseline: PRA ≤ 1 ng/mL/h with PAC ≥ 10 ng/dL.
- Post SST: PRA ≤ 1 ng/mL/h with PAC ≥ 7.8 ng/dL as per the 2025 Endocrine Society Guidelines (20).

▪ Classifications were also repeated using a higher post-SST PAC cutoff (≥10 ng/dL), per 2016 Endocrine Society Guidelines (22).
- Post OSLT: PRA ≤ 1 ng/mL/h with PAC ≥ 10 ng/dL.
- Low-Renin Phenotype: Characterized by low-renin physiology with low concentrations of aldosterone and often referred to as “low-renin hypertension” that could reflect a mild or subclinical phenotype of PA pathophysiology or a non-PA phenotype (23):

- At baseline: PRA ≤ 1 ng/mL/h with PAC <10 ng/dL.
- Post SST: PRA ≤ 1 ng/mL/h with PAC <7.8 ng/dL.
- Post OSLT: PRA ≤ 1 ng/mL/h with PAC <10 ng/dL.

*Renin-Dependent Aldosteronism Phenotype*:

- Characterized by high-renin aldosteronism that steers away from the possibility PA (20) and often implicates renal-vascular disease and/or sympathetic nervous system activity:

- At baseline, Post-SST, and Post-OSLT: PRA> 1 ng/mL/h regardless of PAC.

When a participant retained their baseline phenotype following dynamic testing, it was referred to as “persistent,” whereas if a new phenotype emerged following dynamic testing, it was referred to as “unmasked”.

#### ACTH-Independent Hypercortisolism Phenotype

Participants underwent a DST to assess for an ACTH-independent hypercortisolism phenotype: defined as a morning serum cortisol concentration ≥ 1.8 mcg/dL, along with ACTH <10 pg/mL, following overnight dexamethasone(24).

#### ACTH-Modifiable Aldosterone and Cortisol Production

ACTHstim and DST were used to assess ACTH-modulated aldosterone and cortisol levels within aldosterone phenotypes: The pattern of aldosterone and cortisol production following dexamethasone and ACTHstim was compared among individuals with a PA phenotype, low-renin phenotype, and renin-dependent aldosteronism phenotype.

### Laboratory Assays

PAC was measured using a commercially available ELISA (IBL-America; Minneapolis, MN, USA; RRID: AB_2813725), with an inter-assay coefficient of variation (CV) of 8.6%–9.4%, intra-assay CV of 3.9%–9.7%, analytical sensitivity <5.7 pg/mL, and a dynamic range of 5.7–1000 pg/mL. PRA was measured by ELISA (IBL-America, RRID: AB_3532145), with inter-assay CV of 4.8%–7.1% and intra-assay CV of 6.3%–8.7%.

24-hour urine collections were used to measure aldosterone by ELISA (IBL-America; as previously described) to reflect the 24-hour urinary aldosterone excretion rate. Cortisol was measured using the Access Cortisol chemiluminescent immunoassay (Beckman Coulter, Fullerton, CA, USA; RRID:AB_2802133), which has a dynamic range of 0.4-60.0 µg/dL, with intra-assay CVs of 4.4-6.7% and inter-assay CVs of 6.4-7.9%. ACTH was measured by ELISA (American Laboratory Products Company, NH, USA; RRID:AB_3714847), which has a dynamic range of 6.4-476 pg/mL, with intra-assay CVs of 2.3-6.7% and inter-assay CVs of 6.9-7.1%. Urinary free cortisol (UFC) was measured by liquid chromatography-tandem mass spectrometry (LC-MS/MS) following solid phase extraction. The UFC assay has a lower limit of quantification of 1.0 ng/mL, a linear range of 1-100 ng/mL and intra- and inter-assay coefficients of variation ranging from 3.3-7.7% and 4.3-19.5%, respectively.

### Statistical Analysis

Baseline characteristics are reported as frequencies and percentages for categorical variables and as mean ± standard deviation (SD) or median with interquartile range (25th–75th percentile) for continuous variables, depending on distribution. Spaghetti plots, dot plots, and Sankey diagrams were used to visualize the data depicting intra-individual trajectories and distributions showing the raw data. Group comparisons between post-SST plasma renin groups (≤1 vs >1 ng/mL/h) were assessed using Wilcoxon rank-sum tests for continuous variables and Fisher’s exact tests for categorical variables. Cortisol and aldosterone levels post-DST and post-ACTHstim were compared across phenotypes using one-way ANOVA. When the overall test was significant, Tukey’s post-hoc test was used to identify which phenotype pairs differed. All statistical analyses were conducted using RStudio (Version 2024.12.1+563).

## RESULTS

### Study Population

The mean age was 55.4 ± 9.4 years and 66.2% of participants were women. Most participants were White (83.1%) with a mean BMI of 34.8 ± 5.2 kg/m². The mean screening blood pressure was 128.9 ± 11.2 over 81.9 ± 8.4 mmHg, with 67.5% (52/77) of participants being on antihypertensive medication at screening. Median, seated PRA and PAC on *ad libitum* dietary intake, and following withdrawal of diuretics and renin-angiotensin-aldosterone system inhibitors per protocol, were 1.20 ng/mL/hr [0.50, 2.20] and 18.5 ng/dL [13.9, 25.3], respectively (**Table 1**).

**TABLE 1.**

| Characteristics |  |  |
| --- | --- | --- |
| Study Population, N |  | 77 |
| Age (years), mean (SD) |  | 55.4 (9.4) |
| Female sex, n (%) |  | 51 (66.2) |
| Race, n (%) |  |  |
|  | White | 64 (83.1) |
|  | Black | 11 (14.3) |
|  | Other | 2 (2.6) |
| Body mass index (kg/m<sup>2</sup>), mean (SD) |  | 34.8 (5.2) |
| Antihypertensive Medication, n (%) |  |  |
|  | Diuretics* | 9 (11.7) |
|  | ACE inhibitors/Angiotensin II receptor blockers* | 27 (35.1) |
|  | Calcium Channel Blockers | 13 (16.9) |
|  | Beta-Blockers | 3 (3.9) |
|  | Alpha<sub>2</sub>-adrenergic agonist | 1 (1.3) |
| Screening systolic blood pressure (mmHg), mean (SD) |  | 128.9 (11.2) |
| Screening diastolic blood pressure (mmHg), mean (SD) |  | 81.9 (8.4) |
| 24h systolic blood pressure (mmHg)<sup>#</sup>, mean (SD) |  | 124.2 (8.8) |
| 24h diastolic blood pressure (mmHg)<sup>#</sup>, mean (SD) |  | 73.6 (6.8) |
| Serum potassium (mmol/L), mean (SD) |  | 4.31 (0.36) |
| eGFR (mL/min/1.73m<sup>2</sup>), mean (SD) |  | 88.0 (14.0) |
| Plasma Renin Activity (ng/mL/h)<sup>+</sup>, median [IQR] |  | 1.2 [0.5–2.2] |
| Plasma Aldosterone Concentration (ng/dL)<sup>+</sup>, median [IQR] |  | 18.50 [13.86–25.35] |

\*These antihypertensive medications were withdrawn prior to initiating phenotyping protocols.
<sup>#</sup>Blood pressure measured during medication washout phase during which time a blood pressure of <130/80 mmHg was targeted, if needed, using doxazosin and/or amlodipine.
+ Aldosterone and Renin were measured after medication washout

### Saline Suppression Testing to Phenotype Hormonal Mechanisms of Hypertension in Obesity

We first evaluated renin dynamics. At baseline, on an *ad libitum* dietary sodium intake, 49.4% (38/77) participants had a low renin (plasma renin activity ≤ 1 ng/mL/h).

Following SST, this proportion increased to 76.6% (59/77), indicating volume-mediated renin suppressibility in 21 additional individuals. In contrast, the remaining 23.4% (18/77) of participants exhibited non-suppressible renin despite saline loading during SST, indicating a persistent renin-dependent aldosteronism phenotype (PRA> 1 ng/mL/h both before and after SST) (**Figure 1**). These participants with a persistent renin-dependent phenotype had higher plasma aldosterone concentrations at baseline, and after both sodium loading maneuvers (post-SST and post-OSLT), and exhibited greater kaliuresis, when compared to individuals with suppressible renin (**Table 2**).

**Figure 1:**
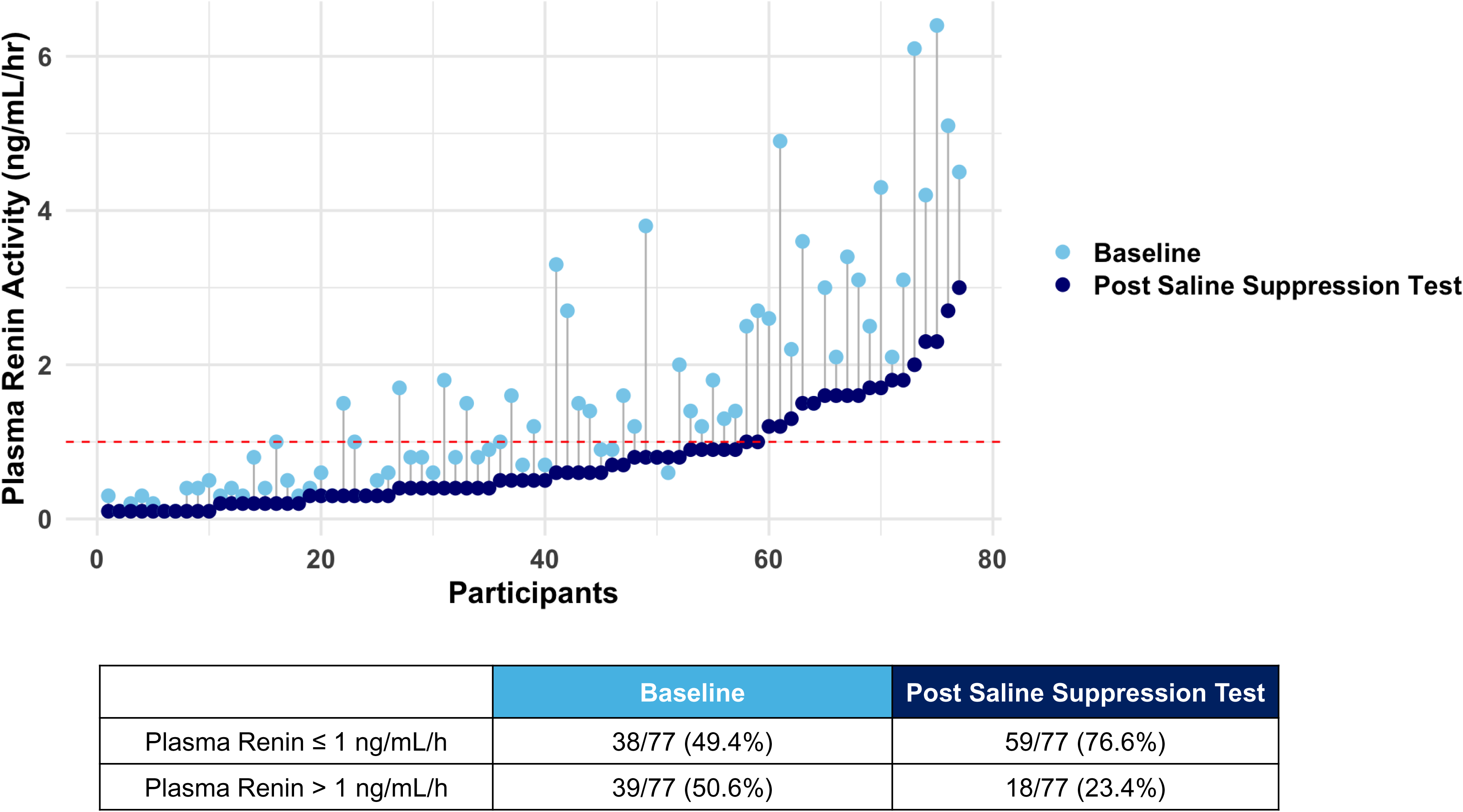
Plasma Renin Activity Before and After Saline Suppression Test. Spaghetti plot representing individual participant’s plasma renin activity at baseline (light blue) and after the saline suppression test (dark blue). The dashed red line indicates the cutoff of 1 ng/mL/hr. At baseline, 38 of 77 participants (49.4%) had plasma renin activity ≤ 1 ng/mL/hr, while 39 of 77 (50.6%) had plasma renin activity> 1 ng/mL/hr. After the saline suppression test, 59 of 77 participants (76.6%) had a suppressed renin ≤ 1 ng/mL/h, while 18 of 77 (23.4%) remained non-suppressed.

**TABLE 2.**

|  | **Post SST**<br>**PRA ≤ 1 (n=59)** | **Post SST**<br>**PRA > 1 (n=18)** | ***p-value*** |
| --- | --- | --- | --- |
| **Female sex, n (%)** | 40 (67.8) | 11 (61.1) | 0.78 |
| **Age (years), mean (SD)** | 55.4 (9.8) | 55.3 (8.1) | 0.61 |
| **Body mass index (kg/m<sup>2</sup>), mean (SD)** | 34.3 (4.1) | 36.6 (7.6) | 0.55 |
| **24h mean systolic blood pressure (mmHg), mean (SD)** | 123.5 (8.7) | 126.2 (9.0) | 0.24 |
| **24h mean diastolic blood pressure (mmHg), mean (SD)** | 73.2 (6.8) | 74.5 (7.0) | 0.56 |
| **Serum potassium (mmol/L), mean (SD)** | 4.30 (0.37) | 4.36 (0.35) | 0.60 |
| **Estimated Glomerular Filtration Rate (mL/min/1.73m<sup>2</sup>), mean (SD)** | 87 (15) | 90 (10) | 0.48 |
| **Hemoglobin A1c (%), mean (SD)** | 5.3 (0.4) | 5.3 (0.3) | 0.78 |
| **24h urinary potassium (mEq/24 h), mean (SD)** | 101 (44) | 145 (100) | 0.032 |
| **24h urinary aldosterone (µg/24h), median [IQR]** | 2.7 [1.8–4.0] | 3.1 [1.6–4.9] | 0.67 |
| **Baseline PAC (ng/dL), median [IQR]** | 17.2 [12.6–23.2] | 23.1 [17.4–39.4] | 0.008 |
| **Post-SST PAC (ng/dL), median [IQR]** | 9.9 [7.1–12.5] | 13.7 [8.5–24.3] | 0.010 |
| **Post-OSLT PAC (ng/dL), median [IQR]** | 13.9 [8.9–18.0] | 24.3 [11.2–37.7] | 0.009 |

SD: standard deviation; IQR: interquartile range; PAC: plasma aldosterone concentration; SST: saline suppression test; OSLT: oral salt loading test.

**Figure 2** illustrates the trajectories of overlapping aldosterone phenotypes following SST. At baseline, 37.7% (29/77) of participants had a PA phenotype and 11.7% (9/77) had a low-renin phenotype. Following the SST, the majority of participants with a baseline PA phenotype failed to appropriately suppress aldosterone, consistent with a persistent PA phenotype (28.5%; 22/77), while a minority exhibited aldosterone suppressibility such that they were re-categorized as having a low-renin phenotype (9.1%; 7/77). However, an additional 18 participants (23.4%) who had a renin-dependent aldosteronism phenotype at baseline suppressed their renin following SST without appropriately suppressing aldosterone, thus unmasking a PA phenotype. This increased the overall prevalence of the PA phenotype to 51.9% (40/77) following SST, wherein nearly half of these individuals were only identified post-SST (**Figure 2**).

**Figure 2:**
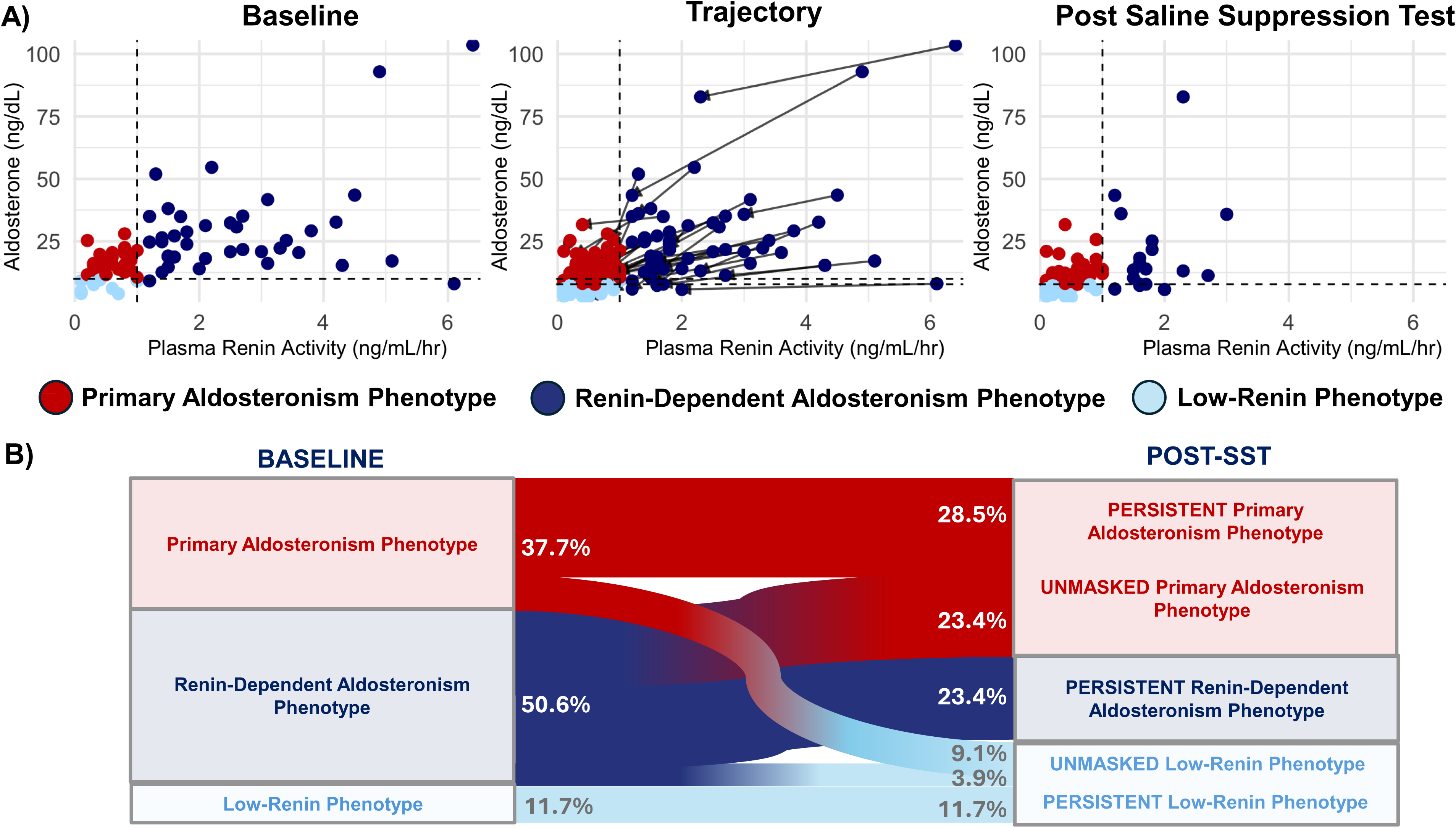
Aldosterone and Renin Phenotype at Baseline and After Saline Suppression Test. **A)** Scatterplots of plasma aldosterone and plasma renin activity at baseline, trajectories from baseline to post saline suppression test, and post saline suppression, showing three phenotypes: (1) primary aldosteronism phenotype in red (PRA ≤1 ng/mL/hr with PAC ≥10 ng/dL at baseline or ≥7.8 ng/dL post-SST), renin-dependent aldosteronism phenotype in dark blue (PRA>1 ng/mL/hr), and low-renin phenotype in light blue (PRA ≤1 with PAC <7.8 ng/dL). **B)** Sankey diagram illustrating phenotype trajectories from baseline to post-saline loading, highlighting persistent (28.5%) and unmasked (23.4%) forms of primary aldosteronism phenotype and persistent renin-dependent aldosteronism (23.4%) phenotype.

These analyses were repeated using the traditional post-SST threshold (≥10 ng/dL), as was the case prior to the recent Endocrine Society guideline update in 2025, yielding similar results, with nearly 40% of participants exhibiting a persistent or unmasked PA phenotype (**Supplemental Figure 1**).

### Validating Hormonal Phenotypes Using Oral Sodium Loading Testing

To validate and replicate the aforementioned findings, a second dynamic test, the OSLT, was performed. Of the 49.2% (32/65) of participants who had a low renin at baseline, this proportion increased to 61.5% (40/65) following OSLT (**Supplemental Figure 2**). 38.5% (25/65) of participants exhibited a persistent renin-dependent aldosteronism phenotype following OSLT (**Supplemental Figure 3**).

In total, 36.9% (24/65) of participants had a PA phenotype at baseline. Following OSLT, the majority of these participants had a persistent PA phenotype (24.6%; 16/65), while a minority exhibited aldosterone suppressibility such that they were re-categorized as having a low-renin phenotype (6/65; 9.2%). However, an additional 9.2% (6/65) participants who had a renin-dependent aldosteronism phenotype at baseline suppressed renin without appropriately suppressing aldosterone post-OSLT, thus unmasking a PA phenotype. This increased the overall prevalence of the PA phenotype to 33.8% (22/65) (**Supplemental Figure 3**).

All 6 (9.2%) participants who demonstrated an unmasked PA phenotype following OSLT also exhibited an unmasked PA phenotype following SST.

### Clinical Implications of Unmasking a PA Phenotype

Of the 51.9% (40/77) of participants who had a PA phenotype following SST (**Figure 2**), less than half (17/40; 42.5%) would have been recognized as having a positive PA screening test at baseline according to Endocrine Society guidelines (PRA ≤ 1 ng/mL/h with PAC ≥ 10 ng/dL and ARR>20) (20). The remainder (23/40; 57.5%) would have been misclassified either due to an elevated renin at baseline (5/23, 21.7%), or because the aldosterone-to-renin ratio was not higher than 20 ng/dL per ng/mL/h (5/23, 21.7%), or both (13/23, 56.5%) (**Figure 3A**). Similar findings were also evident when using the traditional post-SST PAC threshold of 10 ng/dL (**Figure 3B**).

**Figure 3:**
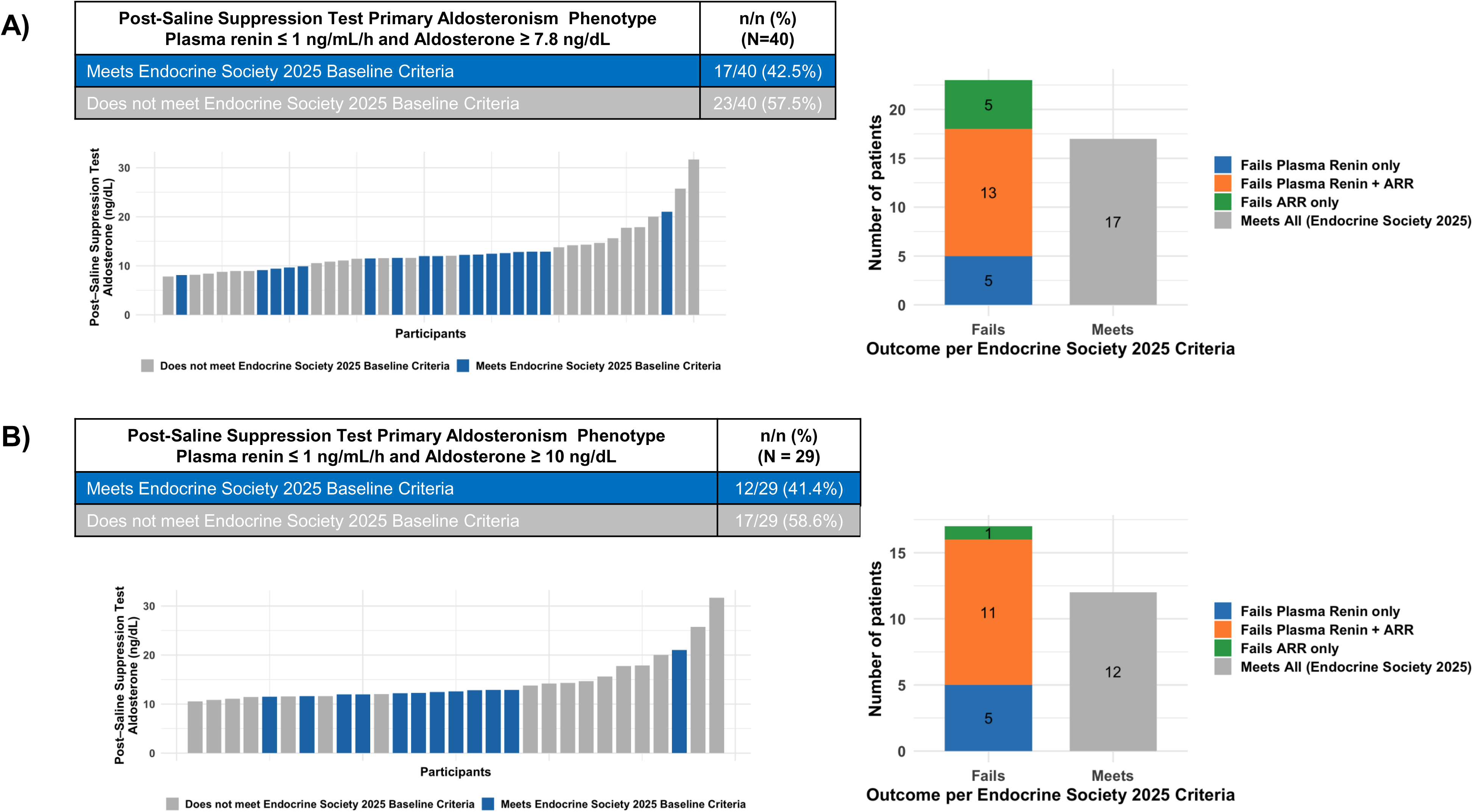
Performance of 2025 Endocrine Society Guidelines in identifying PA phenotype in participants with obesity and hypertension. **A)** Patients with positive saline suppression test (PRA ≤ 1 ng/mL/hr and PAC ≥ 7.8 ng/dL) were identified as having PA phenotype. Among these, only 42.5% would have screened positive at baseline by 2025 Endocrine Society criteria, while the remaining 57.5% failed baseline screening due to non-suppressed renin, low aldosterone-to-renin ratio, or both. **B)** Using the stricter post saline suppression test aldosterone cutoff (≥ 10 ng/dL), 41.4% would have screened positive at baseline, leaving 58.6% missed by Endocrine Society screening, predominantly due to combined non-suppressed renin and low aldosterone-to-renin ratio.

### Cortisol Modulation to Phenotype Hormonal Mechanisms of Hypertension in Obesity

Following DST, 9.2% (7/76) of participants had evidence of ACTH-independent hypercortisolism. All of these participants had normal 24h UFC levels, consistent with findings typically observed in mild autonomous cortisol secretion (MACS) (median 25.3 µg/24 h [IQR 16.1–32.4]) (25). In addition to hypercortisolism, these 7 participants also co-exhibited aldosteronism phenotypes post-SST: PA phenotype in 5/7 participants (71%), of which 3 were unmasked and 2 were persistent, and a low-renin phenotype in 2/7 participants (29%).

### ACTH-modifiable Aldosterone and Cortisol Production to Characterize Established Phenotypes

When examining aldosterone and cortisol concentrations post-DST, cortisol did not differ between phenotypic categories, whereas ACTH-independent aldosterone concentrations were highest among those with a renin-dependent aldosteronism phenotype (*p <0.01*) (**Figure 4A**). Following ACTHstim, stimulated cortisol concentrations did not differ between phenotypic categories; however, ACTH-dependent aldosterone production was highest amongst those with the renin-dependent aldosteronism phenotype (*p <0.001*) (**Figure 4B**).

**Figure 4:**
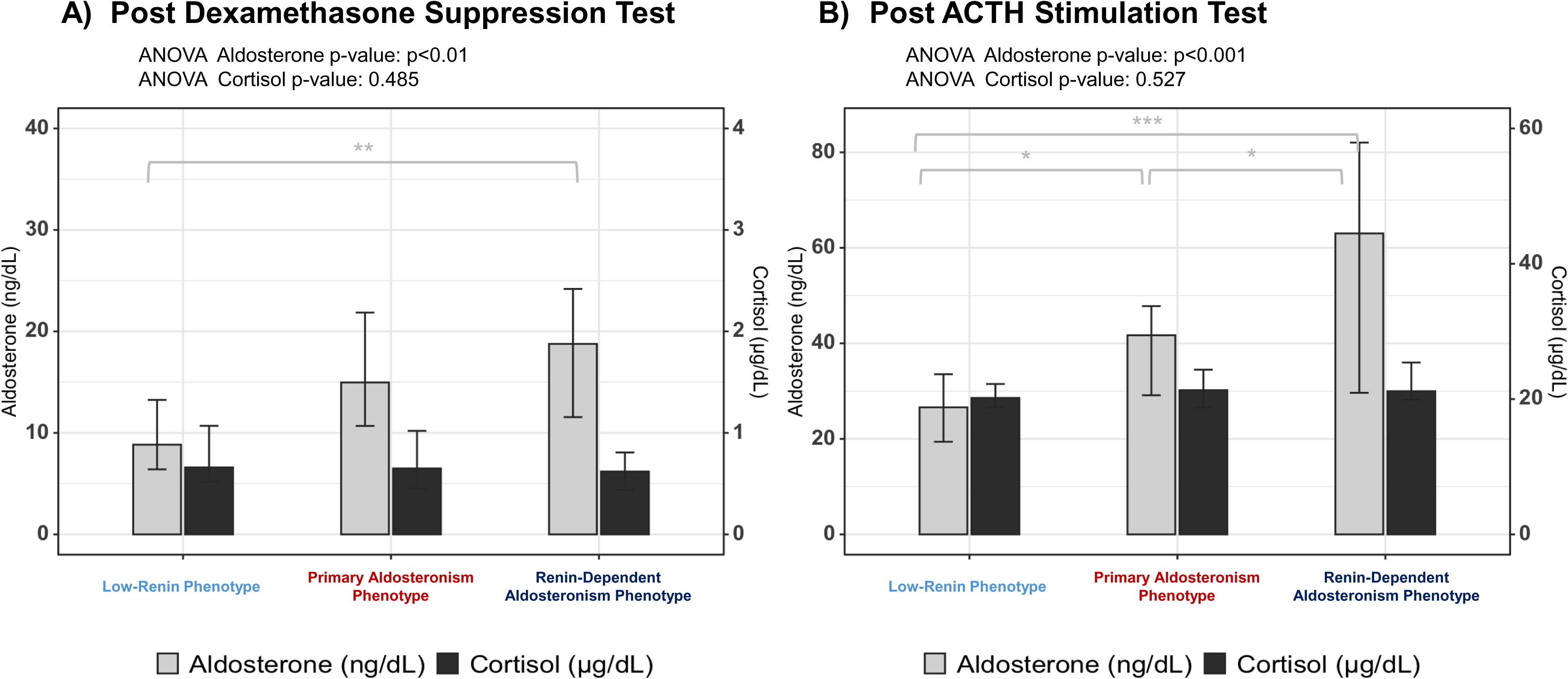
Aldosterone and Cortisol Responses to Dexamethasone Suppression and ACTH Stimulation Test Across Phenotypes. **A)** Post-dexamethasone suppression test: Aldosterone (light gray bars) but not cortisol (dark grey bars) differed significantly across phenotypes (ANOVA *p <0.01* for aldosterone; *p=0.485* for cortisol), with renin-dependent phenotype yielding the highest ACTH-independent aldosterone values. **B)** Post-ACTH stimulation test: Aldosterone showed significant differences between phenotypes (*ANOVA p<0.001*), while cortisol responses were similar (*p=0.527*). Again, with renin-dependent phenotype yielding highest ACTH-dependent aldosterone values.

### Overall Prevalence of Aldosteronism or Hypercortisolism Phenotypes

In total, 84.4% (65/77) of participants had some form of pathologic aldosteronism: 51.9% of participants had a PA phenotype following SST (28.5% with persistent PA and 23.4% with masked PA), another 9.1% had a PA phenotype only at baseline that regressed following SST (potentially representing a milder form of PA or low-renin hypertension), and 23.4% had persistent renin-dependent aldosteronism. Among the 9.2% (7/76) of participants with hypercortisolism, 6.6% (5/76) had an overlapping pathologic aldosteronism phenotype, whereas 2.6% (2/76) did not. Thus, 87% (67/77) of participants with obesity and hypertension had one or more phenotypes of pathologic aldosteronism and/or hypercortisolism.

## DISCUSSION

In this human physiology study, we conducted deep phenotyping studies to interrogate hormonal mechanisms contributing to obesity-related hypertension. Our findings revealed a high rate of overlapping phenotypes of aldosterone production and hypercortisolism. The PA phenotype was identified in over 50% of participants, with approximately 25% exhibiting a clearly recognizable PA phenotype at baseline and another 25% exhibiting a masked PA phenotype that was only revealed following a volume challenge to reveal renin-independent aldosteronism. Importantly, this latter group with masked PA had two concomitant and overlapping phenotypes of aldosteronism: primary (PA phenotype) and secondary (renin-dependent aldosteronism), where the PA component would not have been recognized using conventional screening methods recommended by guidelines (20). Moreover, approximately 25% of participants had persistent renin-dependent aldosteronism where renin could not be suppressed with either intravenous or oral volume loading. These individuals had greater aldosterone production both at baseline and following dynamic maneuvers to modulate ACTH and angiotensin II, and greater kaliuresis, suggestive of greater MR activation. In addition to these pathologic aldosteronism phenotypes, 9% of participants also had clear ACTH-independent hypercortisolism. Taken together, over 80% of participants with obesity and hypertension exhibited one or multiple phenotypes of aldosteronism and/or hypercortisolism.

PA pathophysiology is characterized by renin-independent aldosterone production: inappropriate and relatively non-suppressible aldosterone production when renin is low.

Although we operationalized the evaluation of this PA Phenotype using several well-established interpretations in this study to demonstrate reproducibility and familiarity, it is well known that PA manifests across a continuum of severity(26, 27) such that the magnitude of the renin-independent aldosteronism phenotype is associated with incident hypertension(28–30), cardiovascular disease(31–33), and kidney disease(34). Thus, our categorization of the PA phenotype and low-renin phenotype as distinct entities facilitates the presentation of data in a manner that is familiar to most, though should not be viewed as a literal distinction since the fluidity of renin-independent aldosteronism transcends these categories(26, 27). Our study shows that the phenotype of PA in obesity is more heterogeneous and variable than typically considered, particularly in regard to renin secretion. Greater adiposity has been shown to be associated with overactivation of the RAAS (7, 9, 35), driven by multiple mechanisms, including adipose-derived mineralocorticoid-releasing factors (leptin, angiotensinogen, oxidized fatty acids) and heightened sympathetic activity via altered baroreceptor sensitivity, leptin signaling, and sleep apnea (9, 12, 35–38). Compared with lean individuals, people with obesity have higher circulating levels of angiotensinogen, renin, aldosterone and angiotensin-converting enzyme, all of which are significantly lowered by weight loss (39). Therefore, obesity is regarded as a condition where both renin and aldosterone may be elevated (39). We identified a clear PA phenotype in 37.7% of participants at baseline; however, following saline administration during the SST, renin was further suppressed in an additional 18 participants (23.4%) whose aldosterone production remained inappropriately elevated, thereby unmasking an underlying PA phenotype. Overall, a PA phenotype was present in 51% of participants post-SST, nearly half of whom exhibited overlapping phenotypes of aldosteronism: a renin-dependent phenotype of aldosterone excess at baseline with an underlying masked PA phenotype. Baseline testing was capable of detecting some renin-independent aldosteronism; in contrast, SST induced volume expansion, which lowered renin (40, 41), was required to unmask renin-independent aldosterone production that was not apparent at baseline. Taken together, pathologic phenotypes of aldosteronism, whether primary (renin-independent), or secondary (renin-dependent), or a combination of both, are highly prevalent in individuals with obesity and hypertension and may contribute to cardiorenal disease(19).

Persistent renin-dependent aldosteronism was common, observed in nearly 25% of participants with obesity and hypertension. This phenotype was characterized by higher aldosterone concentrations, greater kaliuresis (signifying greater renal-MR activation), and higher ACTH-modifiable aldosterone levels. ACTH-modifiable aldosterone production is a well-recognized feature of PA, hypothesized to result from altered adrenal sensitivity due to upregulation of the ACTH receptor and somatic mutations within the adrenal cortex (42–44). Here, we demonstrate that a similar ACTH-modifiable phenotype also exists in renin-dependent aldosteronism and appears to be of greater magnitude than that observed in the PA phenotype. A recent study in individuals of metabolically unhealthy obesity showed higher rates of non-suppressible renin and greater salt sensitivity of blood pressure compared to those with metabolically healthy obesity (45). In the context of our findings, the elevation of both AngII- and ACTH-mediated aldosterone, together with greater MR activation, may help explain the more adverse clinical phenotype observed in these individuals.

There is limited data regarding the most efficacious class of medication for treatment for hypertension in obesity. The 2023 European Society of Hypertension Guidelines recommend that patients with obesity be treated with ACE inhibitors, angiotensin II receptor blockers (ARBs), or calcium channel blockers, due to their minimal adverse metabolic effects (8). While ACE inhibitors and ARBs may be effective for the subset of patients with renin-dependent aldosteronism, these agents would not be expected to meaningfully address the more than 50% of participants we observed to have a PA phenotype where aldosterone excess persists even when renin and angiotensin II are low. Given that over 80% of our cohort demonstrated one or more phenotypes of aldosteronism, we postulate that MR antagonists and/or aldosterone synthase inhibitors (ASIs) may represent more effective and mechanistically targeted therapeutic strategies for obesity-related hypertension. Indeed, our findings may help explain the striking blood pressure-lowering effects of ASIs observed in recent trials: in participants with obesity (mean BMI> 30 kg/m²) and resistant or uncontrolled hypertension, ASIs produced near-universal and substantial reductions in blood pressure, suggesting a high prevalence of aldosterone-mediated hypertension (46, 47). In one of these ASI trials (BaxHTN), at least 25% of participants with obesity-related hypertension appeared to have PA based on renin and aldosterone testing at baseline (PRA <1.0 ng/mL/h and aldosterone> 7.5 ng/dL via LC-MS/MS) (47); our current, and prior(26), findings suggest that it is likely that a much larger proportion of the remaining cohort had a masked PA phenotype, milder renin-independent aldosteronism, and/or renin-dependent aldosteronism, thus accounting for the efficacy of baxdrostat. MR antagonists are also hypothesized to be effective for obesity-related hypertension (48–51). Post hoc analyses of the TOPCAT trial found that obese patients with high renin levels experienced the greatest cardiovascular benefit from spironolactone compared with non-obese patients (48, 49), whereas the PATHWAY-2 study demonstrated that the blood pressure-lowering efficacy of spironolactone, especially when renin was low (50, 51).

The dynamics between obesity and hypercortisolism are complex (52). Excess adiposity promotes activation of the hypothalamic-pituitary-adrenal axis, while elevated cortisol further drives adiposity, sodium retention and sympathetic activation (52, 53). Both of these features drive hypertension. In our study, 9% of participants with obesity and hypertension had ACTH-independent hypercortisolism, a comorbidity known to be associated to a higher prevalence of hypertension(54), in part due to cortisol-mediated MR activation.

Our findings should be interpreted in the context of their limitations. First, this study was not designed to be a diagnostic study for PA in obesity. The use of dynamic maneuvers such as the SST, OSLT, DST and ACTHstim are not accurate diagnostic tools for aldosteronism (20, 55, 56); rather, these maneuvers were used in a research setting to interrogate physiology in obesity-related hypertension and ensure reproducible and consistent findings. The interpretations of these dynamic tests have changed over the years; however, regardless of which thresholds are applied to interpret the results of these maneuvers, the underlying findings remain the same: there are multiple, overlapping, and often masked, phenotypes of aldosteronism and hypercortisolism in people with obesity and hypertension. In this regard, the implications of our findings are not to encourage more testing, but rather to understand that nearly all patients with obesity-related hypertension have some manifestation of aldosteronism and/or hypercortisolism that may be amenable to targeted therapy (46, 47, 57). Second, the small sample size limits potential generalizability. On the other hand, this was an unselected cohort recruited from the community that underwent multiple tests and maneuvers that generated highly reproducible findings, thus providing confidence in the mechanisms that were observed. Third, we did not assess treatment responses or clinical outcomes, although recent aldosterone synthase inhibitor trials have already demonstrated effects that bridge our mechanistic findings with established clinical trial evidence (46, 47).

## CONCLUSION

Over 80% of participants with obesity and hypertension exhibited overlapping and pathologic phenotypes of aldosteronism and/or hypercortisolism. The PA phenotype was identified in more than 50% of participants, persistent renin-dependent aldosteronism in nearly 25% of participants, and ACTH-independent hypercortisolism in 9%. These overlapping, and frequently masked, hormonal mechanisms reveal the heterogeneous and multi-factorial nature of obesity-related hypertension and suggest that aldosterone-and cortisol-directed therapies may be the optimal methods to treat blood pressure and abrogating adverse outcomes.

## Data Availability

All data produced in the present study are available upon reasonable request to the authors

## Abbreviations

PA: Primary aldosteronism
RAAS: renin-angiotensin-aldosterone system
BMI: body mass index
MACS: mild autonomous cortisol secretion
MR: mineralocorticoid receptor
MRAs: mineralocorticoid receptor antagonists

---

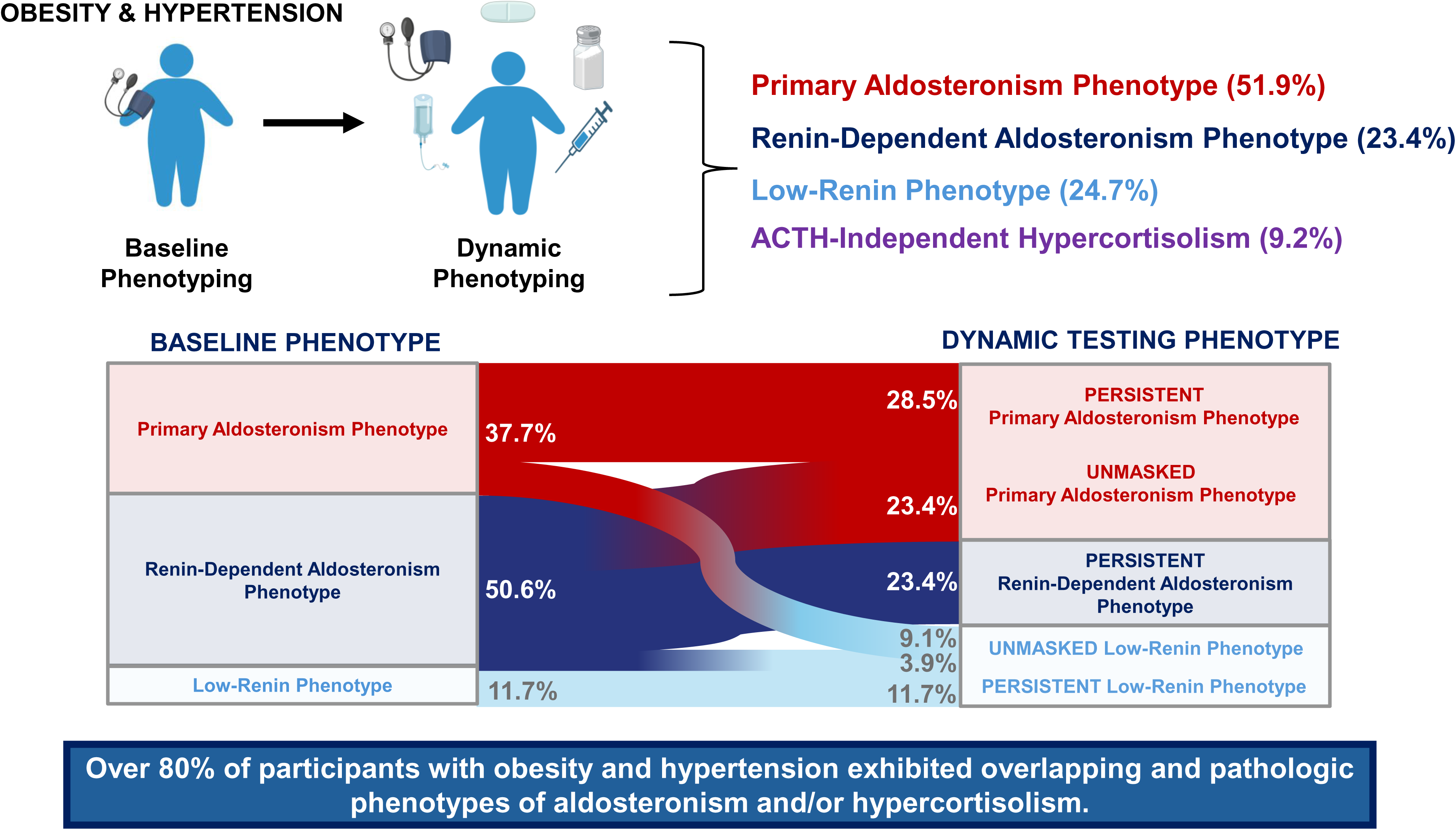

