## Supplementary figures and images for "Unmasking Hormonal Mechanisms of Hypertension in Obesity"

### Supplemental Figure 1

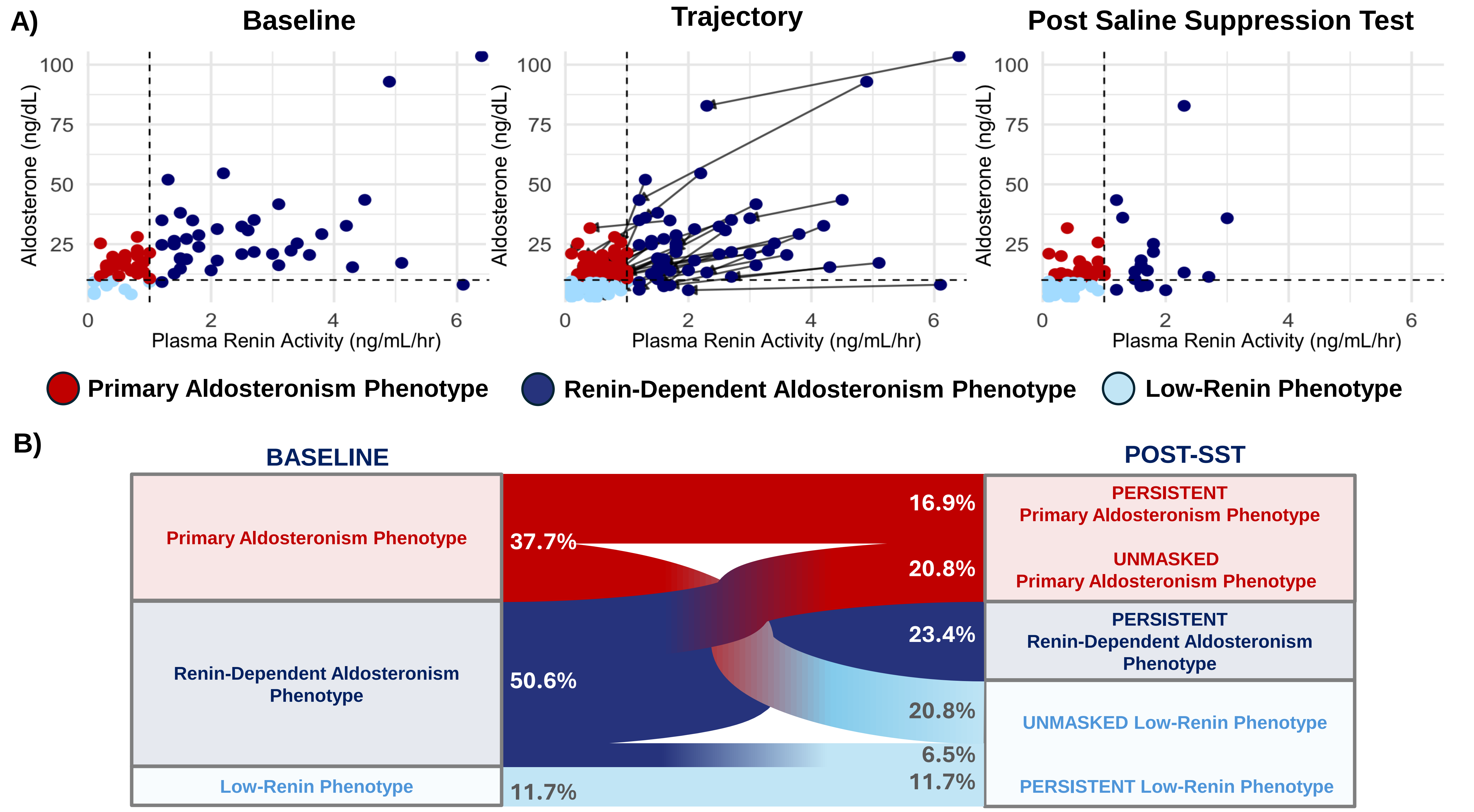

### Supplemental Figure 2

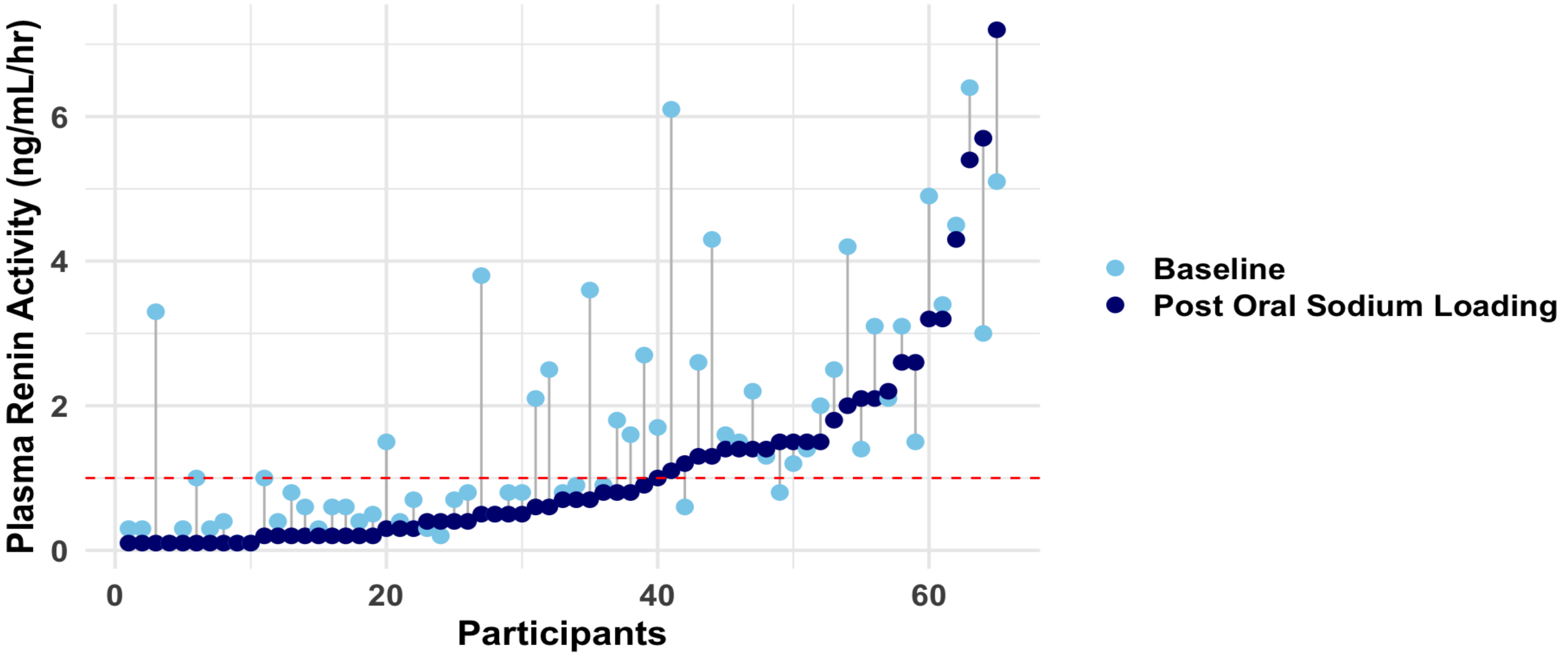


| | Baseline | Post Oral Sodium Loading |
|---|---|---|
| Plasma Renin ≤ 1 ng/mL/hr | 32/65 (49.2%) | 40/65 (61.5%) |
| Plasma Renin > 1 ng/mL/hr | 33/65 (50.8%) | 25/65 (38.5%) |

### Supplemental Figure 3

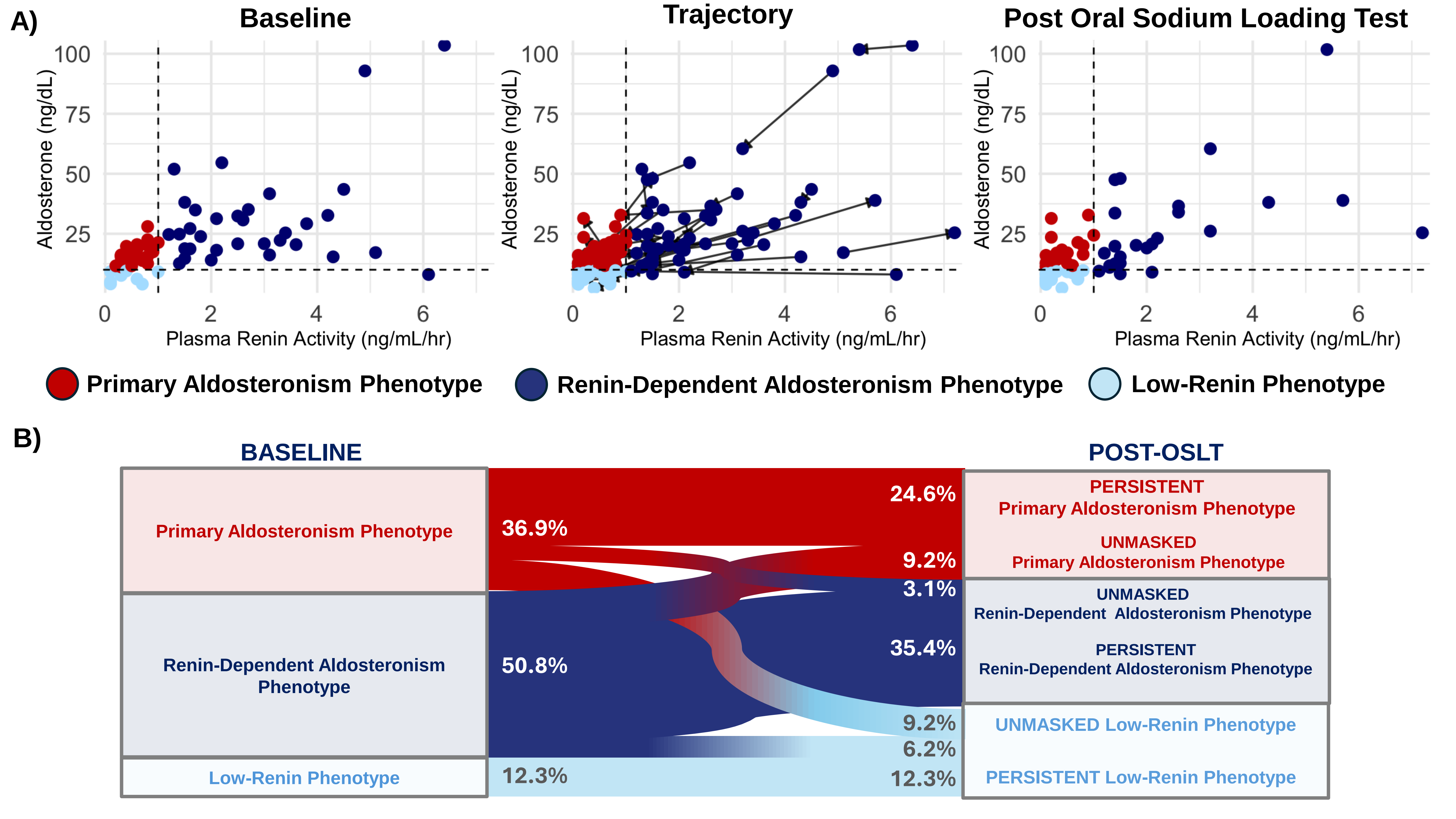
